# Asymptomatic infection in a proof-of-concept longitudinal study on SARS-CoV-2 vaccine recipients

**DOI:** 10.1101/2025.11.26.25341060

**Authors:** Chiaho Shih, You-Zhen Liao, Che-Yu Hsu, Ying-Chen Tsai, Ming-Yen Lin, Hsin-Ying Clair Chiou, Wen-Hui Kuan, Chih-Hsu Chang, An-Ting Liou, Yu-Chi Chou, Feng-Zu Sheen, Hsiang-Jung Lin, Jih-Jin Tsai, Ping-Chang Lin, Ming-Lung Yu, Wan-Long Chuang, Jia-Jung Lee, Jer-Ming Chang, Shang-Jyh Hwang, Justin Shih, Wen-Chun Hung, Ming-Feng Hou, Inn-Wen Chong, Yuh-Jyh Jong, Jung-San Chang

**Author notes:** YZL, CYH, YCT, and MYL made equal contributions to this work. To whom correspondence should be addressed -92.

## Abstract

Individuals with asymptomatic SARS-CoV-2 infection can unknowingly transmit the virus, yet identifying such infections in vaccinated populations remains challenging. We conducted a longitudinal study of 129 vaccine recipients immunized with various combinations of SARS-CoV-2 spike (S) protein vaccine platforms. Sera were collected before the first dose (v1), at 2 weeks (v7) and 6 months (v8) after the third dose. Taiwan’s first major COVID-19 outbreak occurred between v7 and v8. We measured anti-nucleocapsid (anti-N) and anti-S IgG antibody titers by ELISA and assessed virus-neutralizing activity using live virus and pseudovirus assays. By developing an iterative serial screening method, we identified asymptomatic infections among unconfirmed cases. Our v7-v8 paired cohort resolved into three distinct groups: confirmed cases (21%), asymptomatic infections (17%), and uninfected cases (62%). In normalized v8 sera, confirmed cases exhibited an anti-S+++/anti-N+++ phenotype, while uninfected cases showed an anti-S+/anti-N+ phenotype. Statistical analysis validated a distinct asymptomatic group characterized by intermediate anti-S but baseline anti-N antibody levels (anti-S++/anti-N+). This approach enables more accurate estimates of infection prevalence and vaccine efficacy.

## Introduction

SARS-CoV-2 (severe acute respiratory syndrome coronavirus 2) emerged as a global human pathogen in December 2019^1,2^. Despite the availability of vaccines and antivirals, numerous challenges persist in controlling COVID-19 (coronavirus disease 2019)^3^, including vaccine hesitancy, long COVID, breakthrough infections, and the emergence of immune escape and drug-resistant variants^4–7^.

Isolating infected individuals from the community is a critical measure for containing epidemics. However, infected individuals do not always manifest clinically apparent symptoms. People with asymptomatic infections remain a major source of pathogen transmission^8^. In unvaccinated populations, asymptomatic infection can be identified by circulating anti-spike (anti-S) antibody (Ab) but not by anti-nucleocapsid (anti-N) Ab^9–11^. However, in countries with high vaccine coverage, every vaccinated immunocompetent individual is expected to be anti-S positive, regardless of viral infection or symptoms. Therefore, detecting asymptomatic infection in vaccinated populations remains challenging. Diagnosis of asymptomatic infection would be highly valuable for epidemiological practice, enabling more accurate estimates of prevalence rates, vaccine efficacy, and herd immunity.

Neutralizing antibodies (nAb) that recognize the SARS-CoV-2 spike (S) protein are considered the best correlate of protection^12,13^. Spike vaccine-induced nAb decline rapidly, regardless of vaccine platform^14^. Similarly, infection-acquired immunoglobulin G (IgG) to the spike protein and virus-neutralizing activity are also short-lived, declining rapidly after symptom onset^15–17^. Generally, infection-induced nAb exhibit a slower decay rate than vaccine-elicited nAb^18–20^. SARS-CoV-2-specific IgG Ab in symptomatic patients decline more slowly than in asymptomatic infections^21^. In addition to anti-S IgG, anti-N IgG also declines after SARS-CoV-2 infection^22^. Comparisons between anti-N positive and negative cases reveal that waning anti-spike nAb can be boosted by breakthrough infection^23^. Overall, anti-virus Ab decline over time until a new infection is encountered.

Here, using an iterative three-step serial screening method, we identified a distinct group of vaccine recipients with asymptomatic infection. Throughout this report, we use the terms “unreported cases” and “asymptomatic infection” interchangeably. Notably, asymptomatic infections exhibited a unique phenotype characterized by baseline anti-N Ab levels but intermediate anti-S Ab levels. These findings demonstrate that undiagnosed asymptomatic infection can be identified in both unvaccinated and vaccinated populations. Our approach can be applied to estimate prevalence rates, vaccine efficacy, and herd immunity more accurately, supporting more rational public health policy. Early identification of asymptomatic infection should contribute to improved disease control.

## RESULTS

As summarized in the Introduction, it remains unclear whether asymptomatic infection can be identified in vaccinated populations. Both anti-N and anti-S Ab naturally decline over time, regardless of whether they are induced by vaccination or natural infection. To address this issue, we examined anti-N and anti-S Ab titers in a longitudinal cohort of vaccine recipients immunized with combinations of different SARS-CoV-2 spike (S) vaccine platforms (Supplementaary Table S1, Fig. S1, and Methods). The timeline of vaccination and serum collection is outlined in Fig. 1. We focused on 52 self-matched pairs of sera collected at 2 weeks (7th visit, v7) and 6 months (v8) after the 3rd dose vaccination. Between v7 and v8, Taiwan’s first major COVID-19 outbreak occurred. Confirmed cases in Taiwan are officially CDC-registered patients with respiratory symptoms and positive RT-qPCR or rapid diagnostic test results.

**Figure 1.**
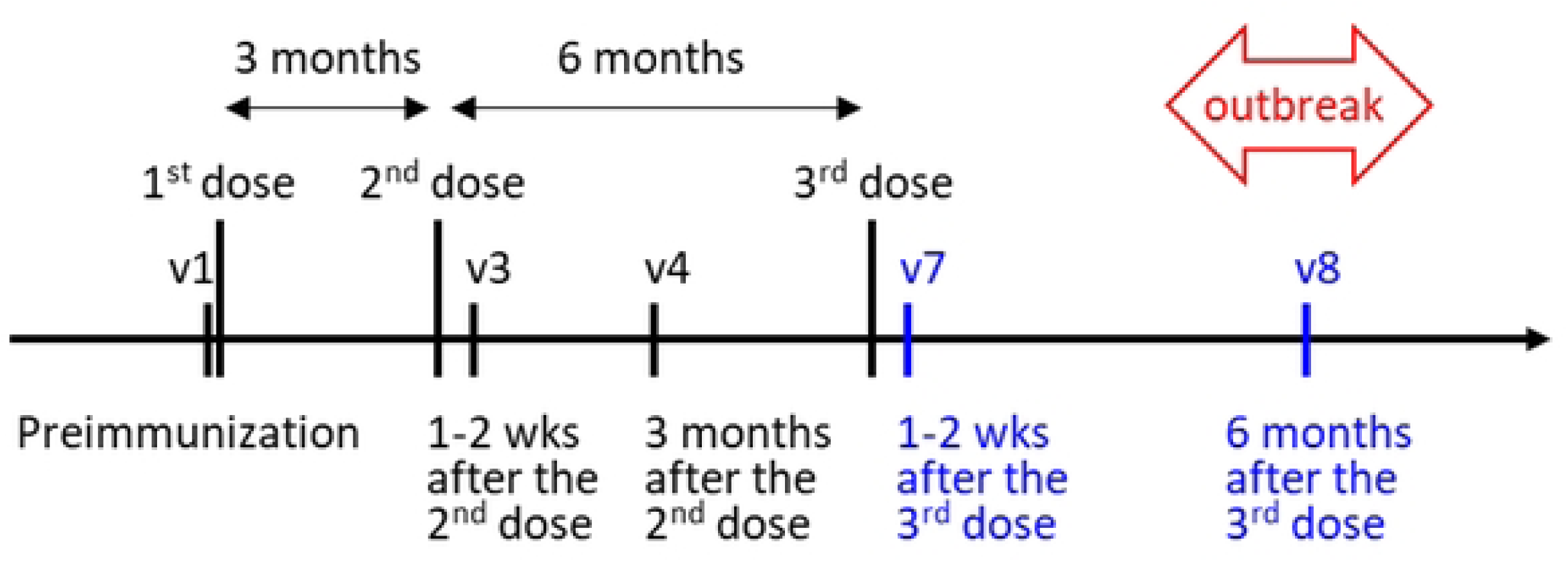
Longitudinal analysis of virus-neutralizing antibody (nAb) in SARS-CoV-2 vaccine recipients. Timeline of participant visits and serum collection at Kaohsiung Medical University Hospital (KMUH). Preimmunization sera were collected at v1 (first visit) one hour before the first vaccine dose in April–May 2021. Participants primed with AZ (AstraZeneca, ChAdOx1 nCoV-19) received a second dose of either AZ three months later or MO (Moderna, mRNA-1273) one to three months later. Serum samples (*v3*) were collected 1–2 weeks after the second dose, and *v4* samples about three months after. A third dose with MO or BNT (Pfizer/BioNTech, BNT162b2) was given six months after the second dose. Some *v4*–*v6* collections were discontinued when the national third-dose campaign began. v7 sera were collected 1–2 weeks after the third dose (December 2021–March 2022), and *v8* sera six months later. A major outbreak occurred between *v7* and *v8* (April–September 2022).

We compared anti-N (nucleocapsid) and anti-S IgG Ab titers between v7 and v8 sera by ELISA and assessed virus-neutralizing activity using live virus and pseudovirus assays. Anti-S Ab titers measured by these assays correlated well with each other (Supplementary Fig. S2). As shown in Supplementary Fig. S3, most sera exhibited a negative slope from v7 to v8 (upper panel, n=52 pairs), indicating waning Ab over this 6-month period. In contrast, confirmed cases exhibited a positive slope, indicating increased anti-S nAb induced by recent breakthrough infection (n=11 pairs, middle panel). Intriguingly, some non-confirmed cases such as A3 and M31 also exhibited a positive slope (bottom panel).

To investigate the cause of positive slopes of anti-S nAb in non-confirmed cases, we plotted anti-S Ab of v7 versus v8 (Fig. 2). The diagonal line indicates near-equal anti-S Ab titers between v7 and v8. Consistent with Supplementary Fig. S3, a subpopulation (orange dots) among non-confirmed cases (n=41; Fig. 2, bottom row) exhibited v8 anti-S Ab titers higher than or equal to v7 samples (v8≥v7). Given the long 6-month interval between v7 and v8, the anti-S nAb of non-confirmed cases should have declined to low levels if no infection occurred during this period (i.e., v7 > v8, blue dots in Fig. 2a-2c, bottom). One potential hypothesis is that these orange dot non-confirmed cases represent asymptomatic infections.

**Figure 2.**
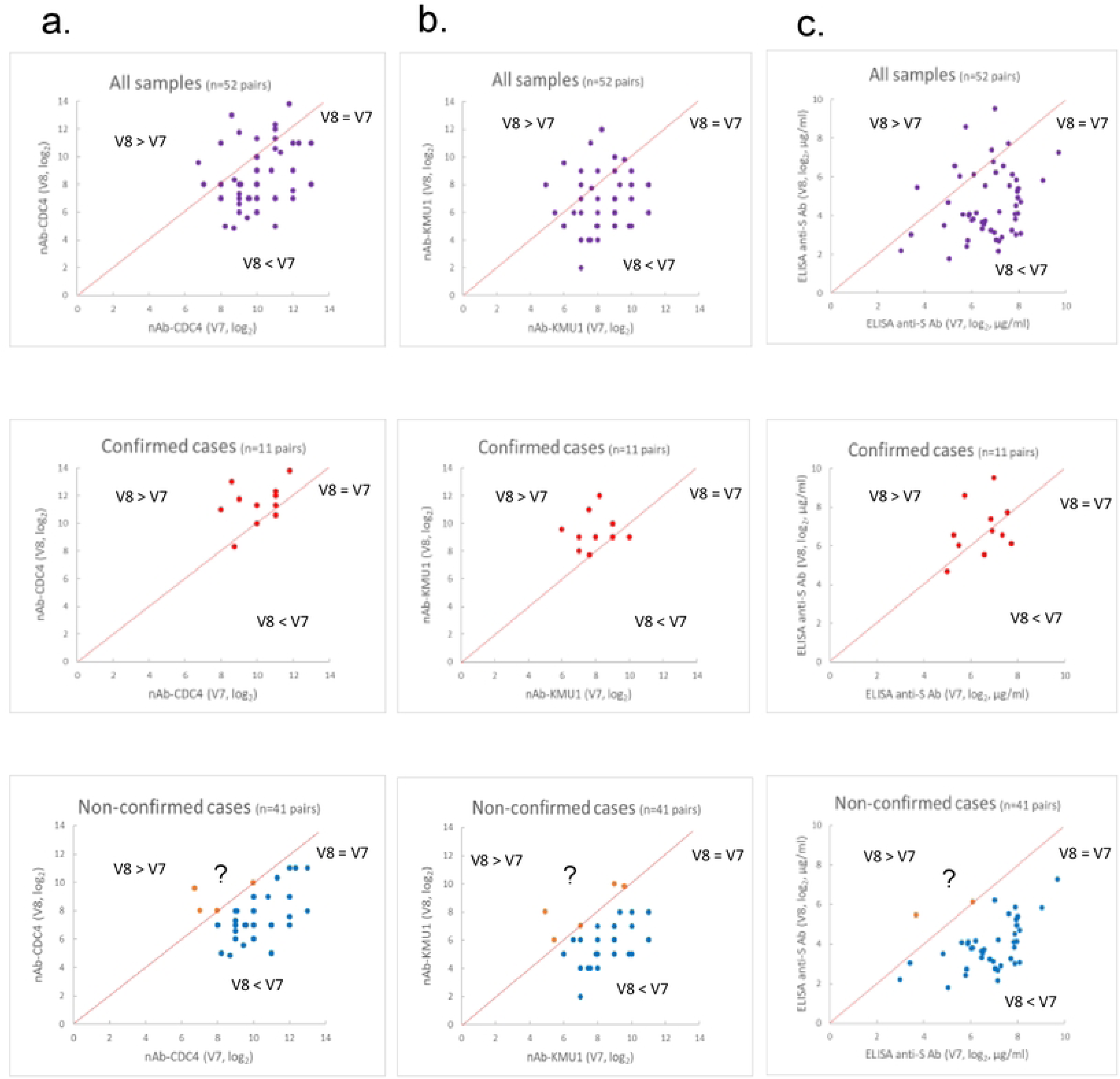
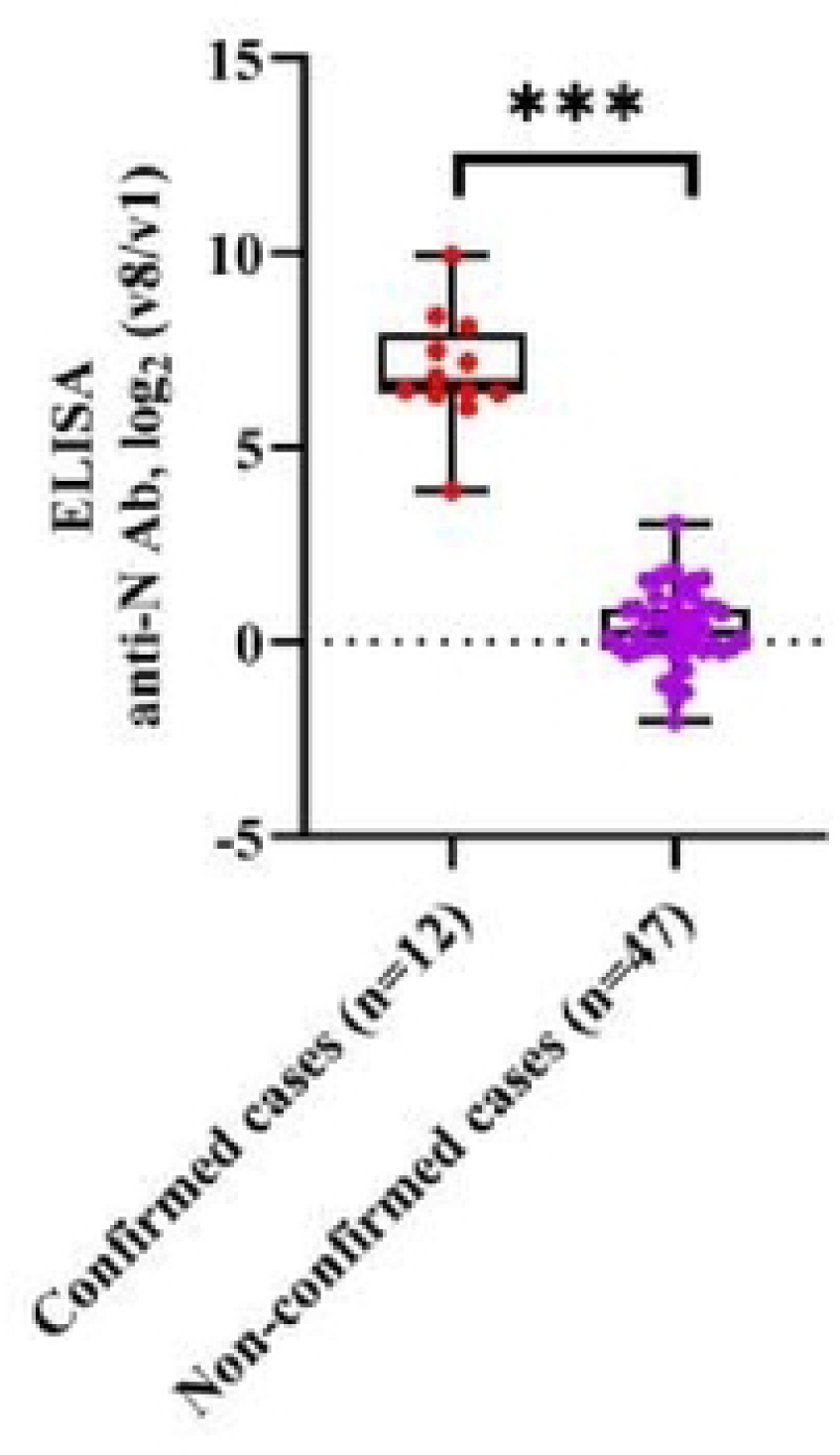
Identification of non-confirmed cases with sustained or elevated anti-S antibody titers. Serum pairs (*v7*, *v8*) from 52 participants were analyzed across assays during the Omicron outbreak (spring 2022). Anti-S antibody titers were measured by (a) live virus neutralization using CDC4, (b) neutralization with the D614G KMU1 variant, and (c) ELISA. Top: all pairs (purple, *n* = 52); middle: confirmed cases (red, *n* = 11); bottom: non-confirmed cases (orange and blue, *n* = 41). Blue dots represent *v8* < *v7*; orange dots denote *v8* ≥ *v7*. Most non-confirmed cases showed declining anti-S antibody titers, though some exhibited unexplained increases (“?”). (d) Confirmed cases (*n* = 12) displayed ∼100-fold higher v8/v1 anti-N antibody ratios than non-confirmed cases (*n* = 41) (two-sided Mann–Whitney U test, ***p < 0.001; median, line; IQR, box). Data represent averages of duplicate measurements from two independent experiments.

Anti-N (nucleocapsid) Ab is a biomarker for SARS-CoV-2 infection^24,25^. We examined ELISA anti-N IgG Ab in all v8 samples (Fig. 2d). Among 13 CDC-registered confirmed cases, the A25 v8 serum is an outlier with 10-fold lower anti-N Ab titer relative to v8 sera from 12 other confirmed cases. This is likely due to earlier v8 sample collection (day 9 after symptom onset, probably before complete viral clearance). Confirmed cases (n=12, excluding the A25 outlier) had an average anti-N Ab of approximately 3.4 mg/ml, which is about 81-fold higher than the 42.3 μg/ml average of non-confirmed cases (n=47). The difference in v1-normalized v8 anti-N Ab (v8/v1) between confirmed and non-confirmed cases was highly statistically significant (p < 0.001; Fig. 2d). In summary, we noted a small group among non-confirmed cases containing v8≥v7 anti-S Ab (Fig. 2a-2c) yet lacking significant anti-N IgG Ab (Fig. 2d).

To systematically define this putative asymptomatic infection group, we designed an iterative 3-step serial screening method to identify candidate asymptomatic infections from non-confirmed cases (Fig. 3a). As shown in Fig. 3b, we plotted a two-dimensional map with a Y-axis of log₂ values of v7-normalized ELISA v8 anti-S Ab titer (v8/v7) versus an X-axis of log₂ values of v1-normalized ELISA v8 anti-N Ab titer (v8/v1). Anti-S IgG Ab naturally declines over time (i.e., v7 > v8) in both spike vaccine recipients and convalescent COVID-19 patients^14–17^. Therefore, candidate asymptomatic infections can be identified from non-confirmed cases if anti-S Ab has v8/v7≥1. However, for non-confirmed cases with anti-S Ab v8/v7 < 1, distinguishing asymptomatic infections from uninfected cases is more difficult. Among all confirmed cases, A82 exhibited the lowest v8/v7 ratio (0.49; Table 1a). We used confirmed case A82 as a primary cutoff (threshold of positivity; dotted line) to screen for candidate asymptomatic infections. Five candidates (A3, A48, A91, M12, and M31) had v8/v7 ratios higher than A82, above the dotted cutoff line (orange dots, Fig. 3b, upper panel). The remaining non-confirmed cases below the dotted line are shown in blue, with several borderline cases labeled with their sample identities.

**Figure 3.**
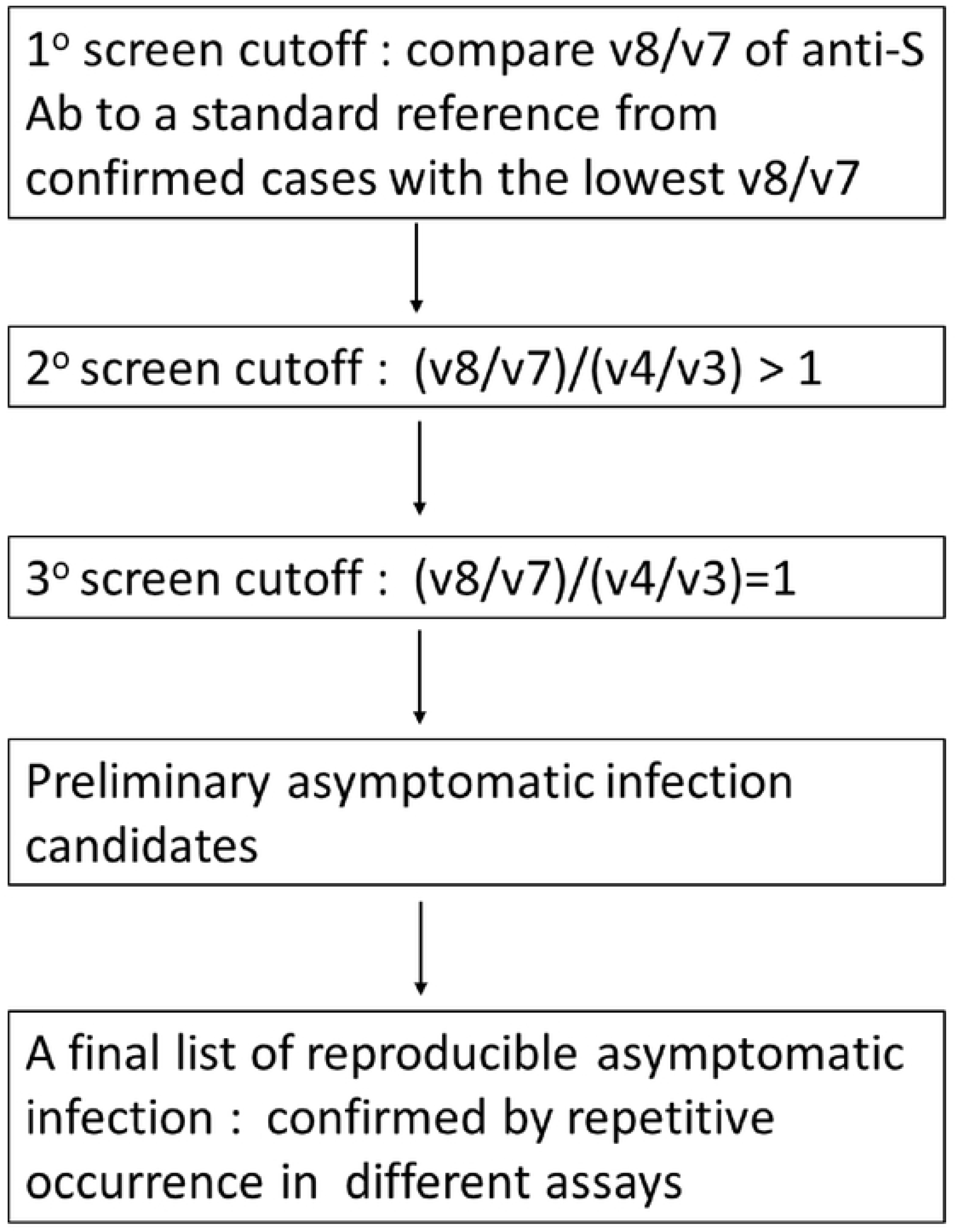
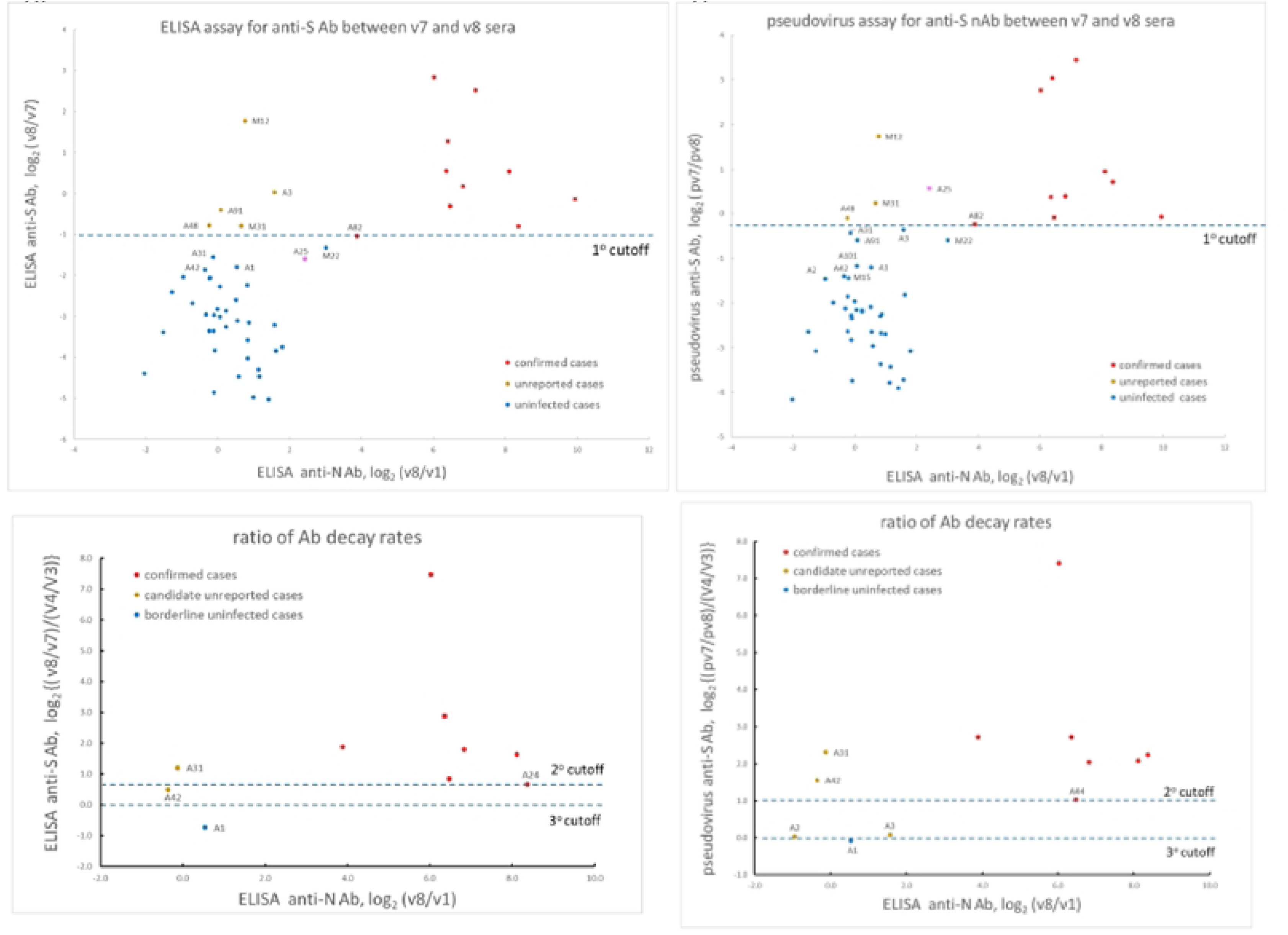
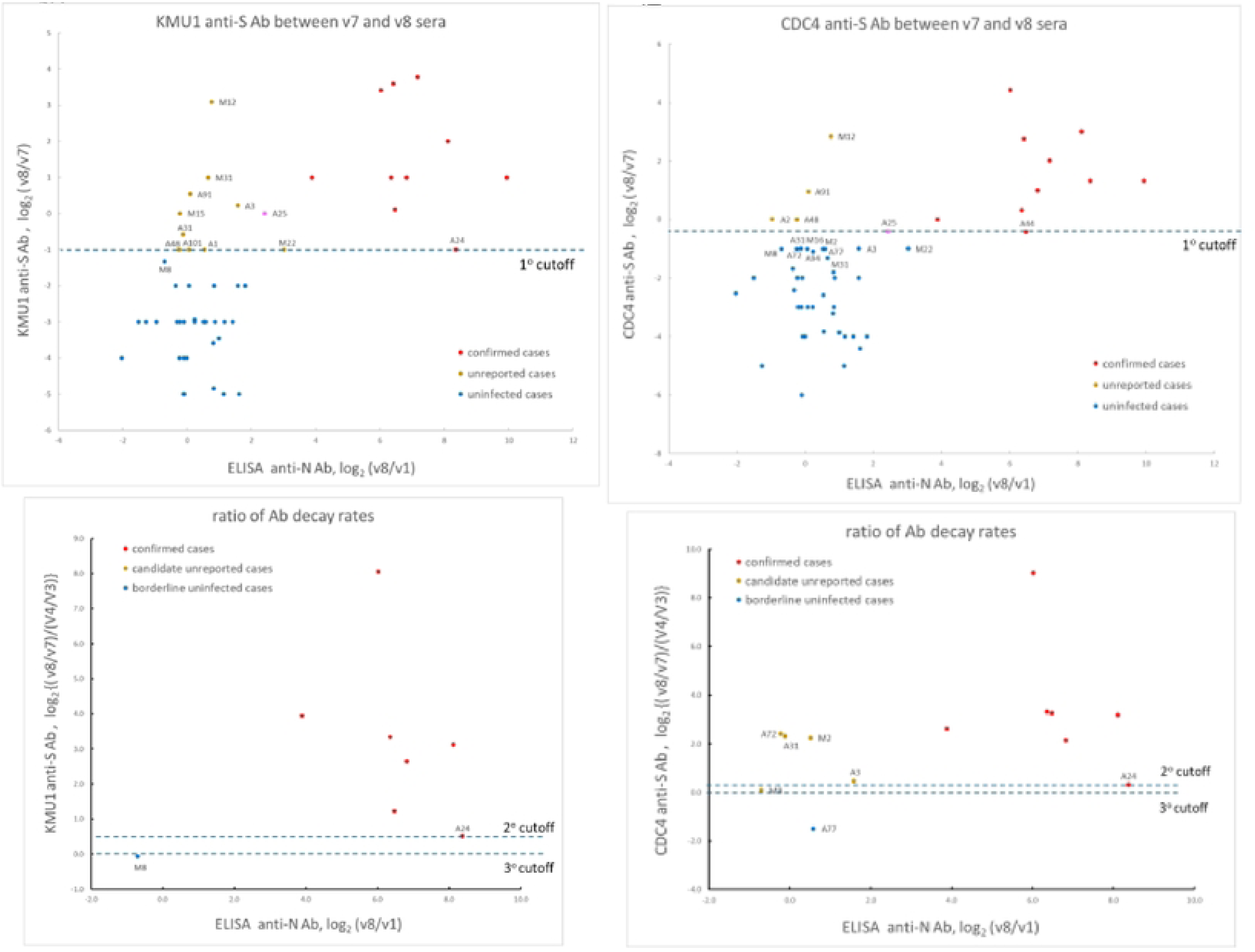
A three-step serial screening strategy for detecting potential asymptomatic SARS-CoV-2 infections. **a**) Overview of the three-step iterative screening method using longitudinal serum pairs (see four examples below). **b) Upper panel:** Double-log plot of ELISA-measured anti-S versus anti-N antibodies in *v8* samples. The *v8*/*v7* log₂ ratio (Y-axis) and *v8*/*v1* log₂ ratio (X-axis) were calculated per participant. Confirmed cases (red) defined cutoff thresholds; A82 (lowest *v8*/*v7*) was used as the primary cutoff (dotted line). Five candidates (yellow: M12, A3, A91, A48, M31) were above the line (Table 1a). Samples below the dotted line are shown in blue. Confirmed case A25 outlier in pink. **Lower panel:** Secondary screening using log₂{(*v8*/*v7*)/(*v4*/*v3*)}; confirmed case A24 provided the cutoff reference. Borderline case A31 crossed the threshold. A tertiary cutoff at log₂{(*v8*/*v7*)/(*v4*/*v3*)} = 0 identified A42 (Table 1b). **c) Upper panel:** Pseudovirus assay comparing anti-S (inverse RLU; *pv7*/*pv8*) to ELISA-measured anti-N antibodies. Confirmed case A82 defined the primary cutoff; A48, M12, and M31 were above it (Table 1a). **Lower panel**: Secondary (A44 cutoff) and tertiary screening (*pv7*/*pv8*)/(*v4*/*v3*) = 1 identified A31, A42, A2, and A3 as candidates (Table 1b). **d) Upper panel:** Live-virus neutralization against KMU1 vs. anti-N ELISA. Using confirmed case A24 as cutoff, ten samples (A1, A3, A31, A48, A91, A101, M12, M15, M22, M31) were identified (Table 1a). **Lower panel**: No additional cases passed secondary/tertiary thresholds. **e) Upper panel:** Live-virus neutralization against CDC4 vs. anti-N ELISA. Samples A2, A48, A91, and M12 exceeded the A44 cutoff (Table 1a). **Lower panel:** Further screening identified A3, A31, A72, M2, and M8 (Table 1b).

**Table 1a.** Preliminary identification of potential asymptomatic infection from the primary screening of a three-step serial screening strategy.

| Assays for anti-S Ab or neutralizing activity | Reference sample used in primary cutoff (v8/v7)* | Candidates of asymptomatic infection with a v8/v7 ratio higher than the primary cutoff | Borderline cases near and below the primary cutoff line** | Reference |
| --- | --- | --- | --- | --- |
| ELISA | A82 (0.49) | A3, A48, A91, M12, M31 | A1, A31, A42, M22 | Fig. 3b, upper panel |
| Pseudovirus*** | A82 (0.85) | A48, M12, M31 | A1, A2, A3, A31, A42, A91, A101, M15, M22 | Fig. 3c, upper panel |
| KMU1 | A24 (0.50) | A1, A3, A31, A48, A91, A101, M12, M15, M22, M31 | M8 | Fig. 3d, upper panel |
| CDC4 | A44 (0.75) | A2, A48, A91, M12 | A3, A31, A72, A77, A94, M2, M8, M16, M22 | Fig. 3e, upper panel |

\* To identify potential asymptomatic infection, we selected a primary cutoff reference from confirmed cases solely based on its lowest value of v8/v7 in each assay. For example, in the ELISA assay, A3, A48, A91, M12, M31 are scored as candidate asymptomatic infections because their respective v8/v7 values are larger than 0.49 of A82, i.e., above the dotted line of the primary cutoff reference A82 in the upper panel of Fig. 3b.
\*\* Borderline cases refer to those non-confirmed cases positioned near and below the primary cutoff line in the upper panels of Fig. 3b-3e.
\*\*\* pseudovirus assay (RLU): pv7/pv8

**Table 1b.** A summary of putative asymptomatic infection by further screening of the borderline case in Table 1a via a secondary and tertiary cutoff.

| Assays for anti-S Ab<br>(v8/v7)/(v4/v3) | Reference sample used<br>in secondary cutoff<br>{(v8/v7)/(v4/v3)} > 1* | Borderline cases with a<br>(v8/v7)/(v4/v3) value higher<br>than the secondary cutoff<br>reference** | Candidates of asymptomatic infection<br>from tertiary screening using a cutoff<br>value of {(v8/v7)/(v4/v3)} = 1 | Reference |
| --- | --- | --- | --- | --- |
| ELISA/ELISA | A24 (1.6) | A31 | A42 | Fig. 3b, lower panel |
| Pseudovirus/ELISA<sup>§</sup> | A44 (2.1) | A31, A42 | A2, A3 | Fig. 3c, lower panel |
| KMU1/ELISA<sup>§</sup> | A24 (1.4) | none | none | Fig. 3d, lower panel |
| CDC4/CDC4 | A24 (1.3) | A3, A31, A72, M2 | M8 | Fig. 3e, lower panel |
| A final list of<br>candidate<br>asymptomatic<br>infection*** | A2, A3, A31, A42, A48, A91, M12, M31, (M15, M22)<sup>#</sup> |  |  |  |

\* The rationale for using (v8/v7)/(v4/v3) as a secondary reference in the screening for potential asymptomatic infection is as detailed in the text.
\*\* New candidate asymptomatic infection from borderline cases in Table 1a were rescreened by using (v8/v7)/(v4/v3) as a secondary cutoff.
\*\*\* These candidates are selected on the final list because they can pass through more than once in different screening assays. For example, samples A1, A72, A101, M2, and M8, are not on this final list, because they can pass through the cutoff only once in only one assay.

To screen for additional potential asymptomatic infections from borderline cases, we used log₂ {(v8/v7)/(v4/v3)} as a secondary cutoff (Fig. 3b, lower panel). While v8 samples were collected 6 months after v7, v4 samples were collected 3 months after v3 (Fig. 1). In theory, the reduction in anti-S Ab after 6 months should be greater than after 3 months. Therefore, for vaccinated individuals with no viral infection experience, the v8/v7 ratio should be less than the v4/v3 ratio. However, v8/v7 could exceed v4/v3 if one or more viral infections occurred between v7 and v8. Since confirmed case A24 had the lowest {(v8/v7)/(v4/v3)} value (1.6; Table 1b), A24 was chosen as the secondary cutoff to screen for additional candidate asymptomatic infections. As shown in the lower panel of Fig. 3b, sample A31 was above the secondary cutoff line. In the third screening cycle, we further relaxed selection stringency by using {(v8/v7)/(v4/v3)} = 1 as a cutoff, and sample A42 (orange) appeared above the dotted line of log₂ 1 = 0 (Table 1b).

Similar to the ELISA assay (Fig. 3b), we repeated the screening using pseudovirus (Fig. 3c) and live virus assays (Fig. 3d, 3e). Table 1 summarizes screening results from these four assays. Many asymptomatic infection candidates selected from one assay were confirmed by a different assay. We compiled a final list of 10 asymptomatic infection candidates: A2, A3, A31, A42, A48, A91, M12, M31, M15, and M22 (Table 1b). The general criterion for inclusion was scoring positive in at least two different assays. Because M15 and M22 lacked v4 serum samples, they could not be screened by the secondary cutoff {(v8/v7)/(v4/v3)}. Nevertheless, based on their overall high v8/v7 values across different assays, M15 and M22 were included in the final asymptomatic infection list. In summary, using an iterative screening method across four different assays, we defined an asymptomatic infection group (n=10) and an uninfected group (n=31) from 41 non-confirmed cases.

Is this newly defined group of unreported cases truly distinct from the other two infection status groups (uninfected and confirmed cases)? As shown in Fig. 4a and Supplementary Table S2, although confirmed cases (red) displayed nearly 100-fold higher anti-N Ab (v8/v1) than the other two groups by statistical analysis (Kruskal-Wallis method with posthoc Dwass, Steel, Critchlow-Fligner test, ***p<0.001), there was no significant difference in normalized anti-N Ab between unreported (orange) and uninfected cases (blue). Next, we compared v8/v7 ratios of anti-S Ab titers between different infection status groups (Fig. 4b and Supplementary Table S2). Both confirmed and unreported cases exhibited significantly higher v8/v7 ratios than uninfected cases in all four assays (***p<0.001). However, significant differences in v8/v7 between confirmed and unreported cases were detected only in the more sensitive pseudovirus assay (**p<0.01) and live virus assay against the ancestral strain CDC4 (*p<0.05), but not in the D614G variant KMU1 or ELISA binding assays. The detected difference in v8/v7 between confirmed and unreported cases in our vaccinated population is supported by a study using an unvaccinated population^21^, showing that symptomatic infection induces higher anti-S-specific neutralizing activity than asymptomatic infection.

**Figure 4.**
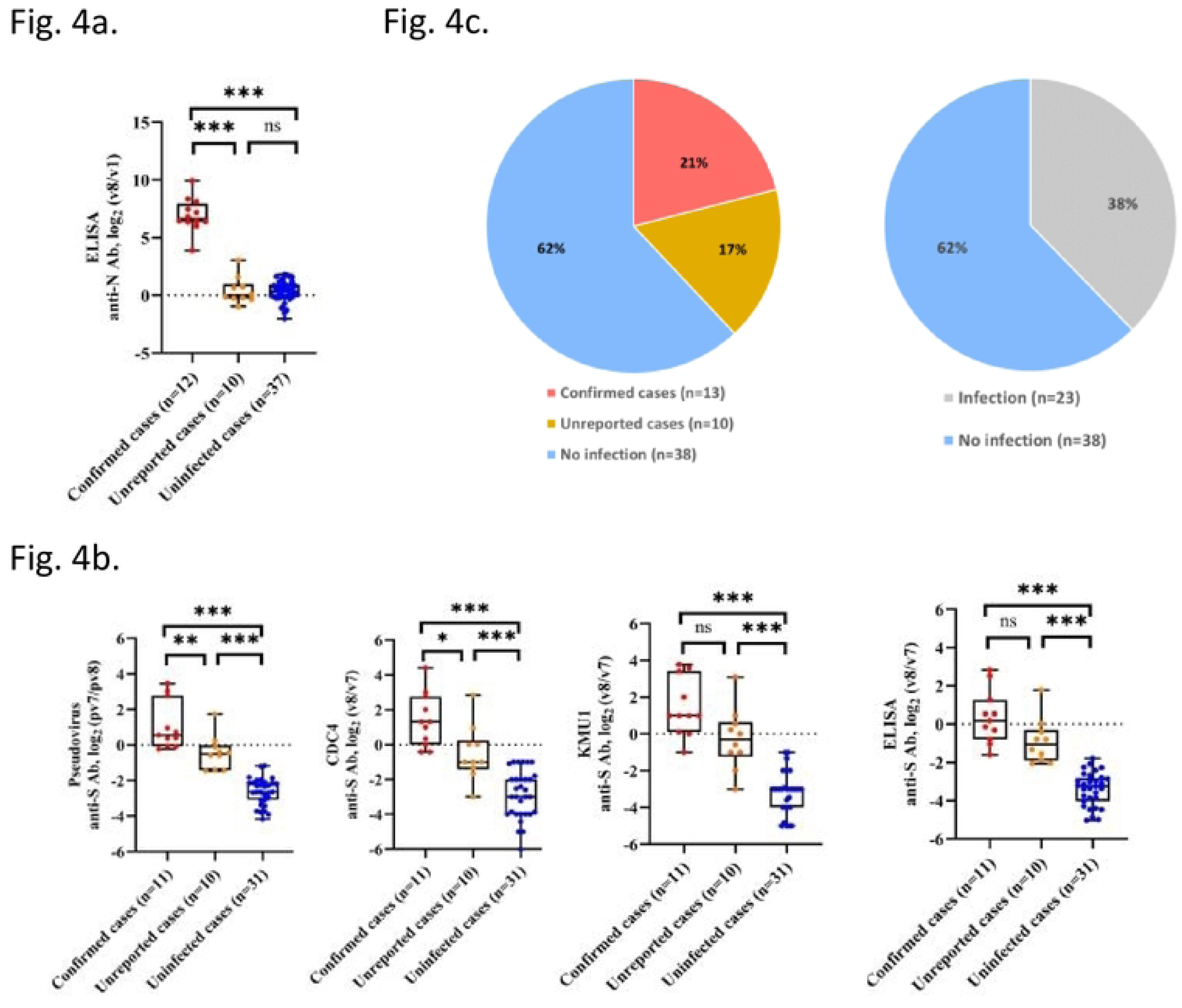
Comparison of antibody titers and infection status distributions in a vaccinated cohort. **a**) Confirmed cases showed significantly higher anti-N antibody levels than unreported or uninfected groups (Kruskal–Wallis with two-sided Dwass–Steel–Critchlow–Fligner post-hoc test, ***p < 0.001). No difference was observed between unreported (*n* = 10) and uninfected (*n* = 37) cases. Dotted line: *v8* = *v1*; median (line), IQR (box). Outliers (A25, A89) were excluded. **b)** Ratios of *v8*/*v7* anti-S antibody titers differed among infection status groups. Confirmed and unreported cases had significantly higher ratios than uninfected cases (***p < 0.001). Significant differences between confirmed and unreported cases appeared in the pseudovirus (**p < 0.01) and CDC4 (*p < 0.05) assays. Dotted line: *v8* = *v7*; median (line), IQR (box). Unpaired confirmed cases (A15, A79) were excluded from ratio analyses (*n* = 11). **c)** Estimated proportions of infection status groups in the vaccinated cohort (*n* = 61): uninfected 62%, confirmed 21%, and unreported 17%. The total infected fraction is approximately 38% (grey). Two unpaired confirmed and seven unpaired uninfected samples were included in the calculation.

As shown in Supplementary Table S2, the median ratios of normalized anti-S Ab (v8/v7 or pv7/pv8) were approximately 10-fold between confirmed and uninfected cases (9.3–19.2), approximately 5-fold between unreported and uninfected cases (3.9–6.4), and approximately 2.5-fold between confirmed and unreported cases (2.1–5). Overall, the difference in normalized anti-S Ab was most significant between infected and uninfected groups.

The pie chart in Fig. 4c summarizes the percentages of confirmed, unreported, and uninfected cases (21%, 17%, and 62%) in our cohort (n=61, including unpaired samples). The total infected population was approximately 38%. None of these three infection status groups showed significant association with age, gender, comorbidities, or vaccine regimens (Fisher’s Exact test, p=0.62) (Supplementary Table S3 and Fig. S4).

## DISCUSSION

In our longitudinal study of vaccine recipients, we designed an iterative screening method to identify putative asymptomatic infections. Asymptomatic candidates exhibited a unique phenotype with intermediate anti-S Ab and baseline anti-N Ab (anti-S++/anti-N+). Our discussion focuses on the potential mechanisms underlying this phenotype and the potential applications and limitations of our approach to detecting asymptomatic infection in well-vaccinated populations.

At least three possibilities could explain the anti-S++/anti-N+ phenotype: 1) An extra spike vaccine dose between v7 and v8. We consider this unlikely since spike vaccine supply was under tight digital control by the government, making it illegal and technically difficult to obtain an extra dose. 2) Technical limits of detection sensitivity of our ELISA kit (Methods). If ELISA detection sensitivity could be further improved, we might detect even lower anti-N Ab levels in some unreported cases. 3) These asymptomatic infection cases experienced transient and abortive infections due to lower-dose virus exposure. Since spike vaccine expresses only spike protein and not N protein, the induction of anti-S nAb in asymptomatic infection cases should be more rapid than anti-N Ab due to three previous spike vaccine doses. Indeed, v8 serum from confirmed case A25 was accidentally collected earlier on day 9 after symptom onset. A25 exhibited high anti-S Ab yet very low anti-N Ab (37 μg/ml average of uninfected cases < 108 μg/ml of A25 < 3413 μg/ml average of confirmed cases). This result is consistent with a previous report showing lower plasma anti-N IgG in vaccinated than unvaccinated patients^25^.

Based on our results in Fig. 4a and Supplementary Table S2, the lack of ELISA-detectable anti-N IgG appears to be a common phenotype in asymptomatic infection in both vaccinated and unvaccinated populations^10,11^. In a South Korean study using unvaccinated individuals, anti-S IgG was detected by ELISA in asymptomatic infections (5/7), but no anti-N IgG was detected by ELISA (0/7)^10^. Similarly, in an unvaccinated Russian cohort^11^, only anti-S Ab was consistently detected, and no anti-N Ab was ever detected in many asymptomatic and mild COVID-19 cases.

Altogether, one explanation for the anti-S++/anti-N+ phenotype of asymptomatic infection is that the exposed S protein on the virion surface may be more immunogenic than N protein in natural infection. Kinetically, anti-N Ab development appears to require a longer time course than anti-S Ab. Unlike confirmed cases, transient SARS-CoV-2 infection during the narrow window of asymptomatic infection is probably insufficient to mount a full-blown N protein-specific humoral immune response.

Preimmunized v1 samples contain pre-existing minor variations in baseline anti-N IgG levels. Therefore, v8 anti-N Ab data required normalization to v1 titers (v8/v1) (Figs. 3 and 4). Similarly, anti-S IgG levels can be strongly influenced by differences in vaccine regimens (Supplementary Fig. S1) and by immunogenetic heterogeneity between individuals in an outbred population. The latter could affect nAb efficacy and durability in the immune response to vaccination. For example, without normalization to its respective v7 counterpart, the neutralizing activity of v8 serum from uninfected sample M8 is even higher than those of unreported cases A2 and A48 (Supplementary Fig. S5). Normalizing the v8 titer to its corresponding v7 titer is an important offset step in our data processing.

Based on v7-v8 paired cohorts from a 3-dose vaccinated population, approximately 17% are candidate asymptomatic infections (Fig. 4c). This 17% estimate is lower than those from unvaccinated cohorts, e.g., 34.9%^26^ and 56%^27^. Clearly, asymptomatic infection rates in vaccinated populations cannot be directly compared with unvaccinated populations. One reason for the difference in estimated percentages of asymptomatic infection could be the more conservative than exhaustive nature of our current study. For example, some residual asymptomatic infections could have cutoff values of {(v8/v7)/(v4/v3)} < 1 in the 3rd screening cycle (Fig. 3). Additionally, some asymptomatic individuals could not meet the criterion of “repetitive occurrence” in two different assays (e.g., A1, A72, A101, M2, M8 in Table 1b). Our current proof-of-concept pilot study collected sera at 3-month (v3-v4) and 6-month intervals (v7-v8). The same principle should be applicable to studies using other flexible interval lengths.

We noted that on rare occasions, results from different assays are not always parallel. For example, v8 sample A81 had nearly 5-fold higher anti-S IgG than v8 sample A84 (34 vs. 7 μg/ml) in the ELISA assay. However, the anti-S nAb titer of A81 was about 2-fold lower than A84 (RLU 1010 vs. 527). The 10-fold discrepancy (2×5) between different assay results does not necessarily imply unreliable assay methods. After all, the Pearson correlation coefficient is very high (r=0.8), though not perfect (r=1.0) (top panel, Supplementary Fig. S2c). We speculate that the neutralizing activity of some samples could involve other unknown immune components, such as anti-S-specific IgA, cytokines, and complement. None of these host factors can be measured by the IgG ELISA assay. Furthermore, the anti-S-RBD protein-based ELISA cannot detect any IgG specific for epitopes outside the RBD.

Vulnerable populations at risk for COVID-19 include healthcare workers and long-term care patients. It might be useful for hospital administration to longitudinally monitor anti-S and anti-N IgG status periodically or during outbreaks. Suspected asymptomatic infections can be promptly self-isolated. In combination with conventional epidemiological tracing techniques, our approach could become a powerful new tool to identify hidden asymptomatic infections.

Another application of our current study is to offer more accurate estimates of prevalence rate, vaccine efficacy, and herd immunity. For example, 21% confirmed cases represent the prevalence rate of symptomatic infection and 17% of our cohort are putative asymptomatic infections (Fig. 4c). Altogether, the prevalence rate of SARS-CoV-2 infection should be 38% (21%+17%). The conventional approach tends to underestimate the overall prevalence rate.

Vaccine efficacy can be evaluated by GMT titer and nAb durability, T cell immune response, or reduction rates of hospitalization and death^28–30^. These assays rely on laboratory settings of tissue culture and animal models or risk differences in disease severity and mortality. Here, we propose a novel alternative indicator for vaccine efficacy using the ratio between numbers of unreported and confirmed cases (e.g., 17% vs. 21% in Fig. 4c). A higher percentage of asymptomatic infection in the total infected population (17%/38%) could suggest more protective vaccine efficacy.

It is possible that lower-dose viral exposure could be more predisposed to asymptomatic infection than higher-dose exposure. While viral dose itself may affect the rate of asymptomatic infection in a pilot study, its influence should be diminished with a larger cohort. Finally, our approach could be applicable to other infectious diseases, including SARS and MERS (Middle East Respiratory Syndrome).

## METHODS

### Study design

During the COVID-19 pandemic, a nationwide vaccination program in Taiwan was initiated in April, 2021. We enrolled a total of 129 SARS-CoV-2 vaccine-naïve participants through Kaohsiung Medical University Hospital (KMUH), Kaohsiung, Taiwan, with the approval of IRB (KMUHIRB-E(I)-20210056). Infection-naïve unvaccinated immunocompetent adults of all ages, genders, and ethnicities were eligible to participate (Supplementary Table S1). Briefly, the mean age of the total 129 participants was 36.2 years. Most were female (70.5%) and without comorbid disease (101/129, 78.3%). Sex was not considered a biological variable in this observational study.

#### Self-matched pairs of samples

These participants received three doses of spike vaccine and were followed up by telephone contact. During this 17-month prospective longitudinal study, an Omicron BA.4/BA.5 outbreak occurred in Taiwan from late April to September, 2022 (Fig. 1). By law, every confirmed case must report to Taiwan CDC and be officially registered. None of our confirmed cases were hospitalized. Antibodies naturally decline over time. Taking advantage of this antibody waning phenomenon, we investigated the issue of asymptomatic infection in a SARS-CoV-2 spike vaccinated population. Loss of serum collection at any stage of follow-up resulted in unpaired samples that could not be included for normalization of antibody titers in the self-matched pairwise analysis. Participants were primed and boosted with various vaccine platforms (Supplementary Fig. S1). Three different three-dose vaccination regimens included AAM (AZ/AZ/MO), AMM (AZ/MO/MO), and MMM (MO/MO/MO). A total of 52 self-matched v7-v8 pairs were available for our study.

### Laboratory Assays

As detailed below, the titer of anti-S Ab was measured by the pseudovirus assay, live virus neutralization against an ancestral viral strains CDC4 and a D614G variant KMU-1, as well as a semi-quantitative ELISA S1-RBD protein binding assay. We observed high to very high Pearson correlations between different assay methods (Supplementary Fig. S2).

#### Cell Lines

A Vero E6-derived cell line was used in live virus neutralization assays. This cell line was established by transfecting the plasmid pDUO2-hACE2-TMPRSS2a (InvivoGen, San Diego, CA, USA) into Vero E6, followed by selecting for hygromycin resistance and stable expression of human ACE2 and TMPRSS2. A human ACE2-expressing HEK293 cell line, designated as ACE2-HEK293 cells, was kindly provided by Dr. Yu-Chi Chou from the RNAi Core of the National Core Facility for Biopharmaceuticals at Academia Sinica, Taipei, Taiwan. The ACE2-HEK293 cells were used in the pseudovirus assay.

#### ELISA

The anti-SARS-CoV-2 N protein Human IgG ELISA kit (Proteintech Co.) has a calibration range 8-128 ng/ml. The anti-SARS-CoV-2 S-RBD (receptor-binding domain) protein Human IgG ELISA kit (Proteintech Co.) has a calibration range 6.25-200 ng/ml. The ELISA is considered a semi-quantitative assay since an internal reference with a serially diluted standard curve was always included in each plate. Each sample was assayed in duplicate in every experiment and an average of two independent experiments was performed for each sample following the vendor’s protocol.

#### Live virus neutralization

Two-fold serially diluted serum samples from vaccinees were examined for live virus-neutralizing activity in 96-well plates using a SARS-CoV-2 ancestral strain CDC4 (hCoV-19/Taiwan/4/2020; EPI_ISL_411927) and the variant strain KMU-1 (hCoV-19/Taiwan/KMU-1/2020; EPI_ISL_4169856, Clade GH, type VI). The KMU-1 spike gene contains only one single D614G mutation. The live virus neutralization assay was conducted in the biosafety level 3 facility at Kaohsiung Medical University Hospital following the approved protocol KMUHIRB-E(I)-20200013. Briefly, 50μl of serially diluted serum samples were loaded in each well of a 96-well plate. In the BSL-III facility, an equal volume (50μl) of CDC4 or KMU1 virus-containing DMEM (10% FBS) (40 TCID_50_/10^4^ cells/well) was mixed with the human serum samples and incubated at 37°C for 1 hour. The mixture of virus-human sera was then transferred to a 96-well plate pre-seeded 16-24 hours earlier with 100μl DMEM (10% FBS) of Vero E6 cells expressing ACE2-TMPRSS2. The culture was incubated in the BSL-III incubator at 37°C for 48-72 hours, followed by fixation with 4% formaldehyde, staining with 0.5% crystal violet, and de-staining with 5% sodium hypochlorite. The titer of neutralizing activity of serum samples was measured by the fold of maximal dilutions that could still protect Vero E6 cells from lysis.

#### Pseudovirus assay

The pseudotyped alpha virus B.1.1.7 was provided by Dr. Yu-Chi Chou at the Biomedical Translation Research Center, Academia Sinica, Taiwan (https://biotrec.sinica.edu.tw/pages/3473). This variant B.1.1.7 contains the following spike mutations, including 69-70 del, Y144 del, N501Y, A570D, D614G, P681H, T716I, S982A, D1118H. Briefly, heat-inactivated human sera were serially diluted to the desired dilution and incubated with SARS-CoV-2 pseudotyped lentivirus in DMEM (10% FBS) for 1 hour at 37°C. The reporter virus titer for a positive control well (no serum added) was typically adjusted to approximately 10,000-20,000 RLU/well/96-well plate. The mixture was then inoculated with 10,000 HEK-293T cells stably expressing human ACE2. Infected cells were continuously cultured for another 48 hours before performing the luciferase assay. The expression level of luciferase was determined using ONE-Glo™ Luciferase Assay System (Promega). The relative luminescence unit (RLU) was detected by SpectraMax Mini. The neutralizing activity (predominantly anti-spike IgG) in the sera is inversely proportional to the RLU value. Each sample was assayed in duplicate in each experiment, and an average of two independent experiments was performed for each sample.

## Statistics

Box plots displayed distributions of anti-S and anti-N antibody titers across three COVID-19 infection status groups. Pearson correlation evaluated ELISA, live virus neutralization, and pseudovirus assays, with coefficients and 95% confidence intervals (CIs). Data were summarized as medians with interquartile ranges (IQRs). Mann-Whitney U compared ELISA anti-N titers between confirmed and non-confirmed cases. Fisher’s exact test, one-way ANOVA, and Kruskal-Wallis test with post-hoc analysis compared participants’ characteristics, vaccination regimens, and antibody titers among groups. Two-sided P-values <0.05 were considered statistically significant.

## Data availability

All data supporting the results of this study are available in this manuscript.

## Acknowledgments

We thank Taiwan CDC for sharing the CDC4 virus. We thank the Biosafety Level-III Core Facility at KMUH for assistance in this study.

## Author contributions

Designed the experiments: CS, JSC; Performed the experiments: CS, YCT, ATL, YZL, WHK, CHC, CYH; Recruitment of participants: IWC, HJL, SJH, JMC, JJL, FZS, YJJ, MLY, WLC, JSC. Provided reagents: HYCC, WCH, MFH, and YCC. BSL-III facility: PCL and JJT. Wrote the paper: CS, CYH, YZL, JS, MYL, JSC. All authors analyzed the data and contributed to discussions.

## Funding

We thank the funding support from National Science and Technology Council, Taiwan (NSTC 113-2320-B-037-030).

## Competing interests

All authors have declared that no conflict of interest exists.

## Additional information

### Supplementary information

The on-line version contains five Supplementary Figures (Fig. S1-S5), and three Supplementary Tables (Table S1-S3).

## Notes

### Funding Statement

Yes

### Author Declarations

We enrolled SARS-CoV-2 vaccine naïve participants through the Kaohsiung Medical University Hospital (KMUH), Kaohsiung, Taiwan, with the approval of IRB (KMUHIRB-E(I)-20210056).

